# Expert panel as reference standard procedure in diagnostic accuracy studies: a systematic scoping review and methodological guidance

**DOI:** 10.1101/2024.11.12.24317219

**Authors:** Bas Kellerhuis, Kevin Jenniskens, Mike P.T. Kusters, Ewoud Schuit, Lotty Hooft, Karel GM Moons, Johannes B. Reitsma

## Abstract

**Background:** In diagnostic accuracy studies, when no reference standard test is available, a group of experts, combined in an expert panel, is often used to assess the presence of the target condition using multiple relevant pieces of patient information. Based on the expert panel’s judgment, the accuracy of a test or model can be determined. Methodological choices in design and analysis of the expert panel procedure have been shown to vary considerably between studies as well as the quality of reporting.

**Objectives:** To map the current landscape of expert panels used as reference standard in diagnostic accuracy or model studies.

**Design:** PubMed was systematically searched for eligible studies published between June 1, 2012, and October 1, 2022. Data extraction was performed by one author and, in cases of doubt, checked by another author. Study characteristics, expert panel characteristics, and expert panel methodology were extracted.

**Eligibility criteria:** Articles were included if the diagnostic accuracy of an index test or diagnostic model was assessed using an expert panel as reference standard and the study was reported in English, Dutch, or German.

**Results:** After initial identification of 4,078 studies, 318 were included for data extraction. Expert panels were used across numerous medical domains, of which oncology was the most common (20%). The number of experts judging the presence of the target condition in each patient was 2 or less in 29%, 3 or 4 in 55%, and 5 or more in 16% of the 318 studies. Expert panel types used were an independent panel (i.e., each expert returns a judgement without conferring with other experts in the panel) in 33% of studies, a panel using a consensus method (i.e., each case was discussed by the expert panel) in 27%, a staged (i.e., each expert independently returns a judgement and discordant cases were discussed in a consensus meeting) target condition assessment approach in 11%, and a tiebreaker (i.e., each expert independently returns a judgement and discordant cases were assessed by another expert) in 8%. The exact expert panel decision approach was unclear or not reported in 21% of studies. In 5% of studies, information about remaining uncertainty in experts about the target condition presence or absence was collected for each participant.

**Conclusions:** There is large heterogeneity in the composition of expert panels and the way that expert panels are used as reference standard in diagnostic research. Key methodological characteristics of expert panels are frequently not reported, making it difficult to replicate or reproduce results, and potentially masking biasing factors. There is a clear need for more guidance on how to perform an expert panel procedure and specific extensions of the STARD and TRIPOD reporting guidelines when using an expert panel.

**Strengths and limitations of this study:**

- This review provides an overview of trends in the use of expert panels as reference standard in diagnostic accuracy studies.
- This review touches on several aspects of expert panel use that have previously not been considered, including incorporation, differential verification, uncertainty in expert judgements and diagnosis using AI.
- Though this review has systematically searched PubMed, other electronic databases have not been searched, so it is possible not all diagnostic accuracy studies using an expert panel as reference standard are included.

## Introduction

Diagnostic accuracy studies evaluate whether a diagnostic test or model under study, also referred to as the diagnostic index test or model, can accurately assess or predict the presence or absence of a target condition, as determined by a reference standard [1]. Diagnostic accuracy measures, such as c-index, sensitivity, specificity, posterior probabilities or predictive values and calibration plots, can be calculated by comparing the results from the index test or model and the reference standard.

When no single reference standard test determining the presence or absence of the target condition is available, an expert panel is often used to assess this in each study patient [2–5]. Such an expert panel typically consists of a group of medical experts in the domain of the target condition of interest, such as medical specialists, nurses, specialized lab technicians, or experienced patient representatives. The expert panel determines the presence or absence of the target condition typically based on multiple relevant pieces of information (e.g., medical history, (biomarker)test results, medical imaging data, follow-up data) documented of the study patient. The final diagnosis by the expert panel can then be used to calculate measures of diagnostic accuracy of the index test or model, or to compare diagnostic accuracy relative to other tests or models.

In 2012, a review outlined the various properties of expert panels used as a reference standard in DTA studies, including the different methods used to operationalize the decision-making process [6]. It revealed that methodological choices in design and analysis of the expert panel procedure varied considerably between studies, and that 83% of studies missed one or more pieces of critical information about the applied methodology, e.g., the number of experts in the panel or the methodology by which a decision is made. Since its publication, new methodological guidance has been published that may have impacted the properties, use, and method of decision-making in expert panels in diagnostic accuracy studies [7].

A recent methodological insight has been published highlighting that forcing a dichotomous classification by the expert panel (i.e., target condition present or absent) may lead to problems. This paper shows that a forced dichotomization ignores remaining uncertainty on the decided presence or absence of the target condition and may lead to biased diagnostic accuracy estimates [8].

Artificial intelligence is increasingly used as a part of diagnostic tests or models. Expert panel assessments frequently play a role in the training and evaluation of artificial intelligence algorithms outside of biomedical research. New techniques tested in other research domains may lead to new opportunities for the set-up of expert panels in diagnostic research. For example, it may be possible that a large group of non-specialists can be used as a substitute for a small group of specialists as an expert panel for some target conditions, allowing for more efficient and less costly diagnostic research [9].

Given these developments, we aimed to update and extend a previous review [6] on the use of expert panels in diagnostic test or model accuracy research to evaluate the variety in the use of and the decision methodology applied in expert panels.

## Methods

For this systematic scoping review, we searched the PubMed database for primary diagnostic test or model accuracy studies that used an expert panel as a reference standard. We aimed to assess contemporary use of expert panels by updating the search by Bertens et al. [6], therefore using 1^st^ of June 2012 as our starting date and 1^st^ of October 2022 as our end date for screening and study selection.

The search string included terminology related to expert panels, consensus diagnosis, diagnostic accuracy, and several common diagnostic accuracy measures, such as sensitivity, specificity, c-index and predictive values. An information specialist assisted in constructing the search strategy. The full search string can be found in appendix A.

### Inclusion and exclusion criteria

Articles were included if the accuracy of a diagnostic index test or model was assessed using an expert panel as a reference standard and the study was reported in English, Dutch, or German. No restrictions were enforced regarding the medical domain, context, setting, or composition of the panel. Articles were excluded if the full text was not available or if they involved non-human subjects (i.e., animals).

Records were screened for eligibility based on title and abstract and thereafter based on full text by one author and checked by the other (BK or MK).

### Data extraction

Data extraction was piloted using a random sample of 20 eligible articles and discussed and improved by the author team to obtain the final set of data extraction items. The data extraction form is presented in appendix B. Data extraction of all included articles was performed by one author (BK). In cases of doubt, a second reviewer (MK) was consulted.

We extracted general study characteristics, expert panel characteristics, and expert panel methodology used for evaluation and decision-making on the target condition presence. General study characteristics included the number of participants in the study, type of index tests or models, whether the index test or model was a machine learning, AI, or software tool, and the target condition assessed in the study.

Expert panel characteristics and methodology included the number of experts in the study, the number of experts in each panel, the expertise of experts included in the panel (as described by the authors), types of information provided to the expert panel (e.g., medical history, biomarkers, imaging, and follow-up data), whether the index test result was incorporated in the information provided to the panel, whether data was pre-labeled (i.e., for a databank or previous study) and whether all participants were assessed in the same way (i.e., whether the expert panel assessed all participants or whether some participants were assessed through another method such as a biopsy).

Expert panel methodology was first assessed by identifying the structure and target condition decision-making process, aiming to identify key variations in expert panel procedures. After this data collection, we constructed 4 main types of expert panels procedures based on whether experts provided independent assessments or consensus was sought from the start and how disagreements were resolved.

Additionally, we assessed whether experts were asked to record their level of certainty about the presence or absence of the target condition for individual patients.

### Statistical analysis

Results were summarized descriptively using percentages for dichotomous and categorical results. Continuous results were summarized using the median and interquartile range and illustrated graphically using histograms. We aimed to identify and define distinct types of expert panel procedures according to their composition and decision-making process. Where of interest, results were stratified by this expert panel type.

This manuscript has been reported according to the Preferred Reporting Items for Systematic reviews and Meta-analyses Extension for Scoping Reviews (PRISMA-ScR) [10]. We did not register a review protocol.

## Results

A total of 4078 studies were identified in our PubMed search. During title and abstract screening 3524 studies were excluded. After removal of 32 studies for which no full text paper was available, another 204 were excluded during full text screening, leaving 318 articles for data extraction and further analysis. A flowchart is presented in Figure 1.

**Figure 1:**
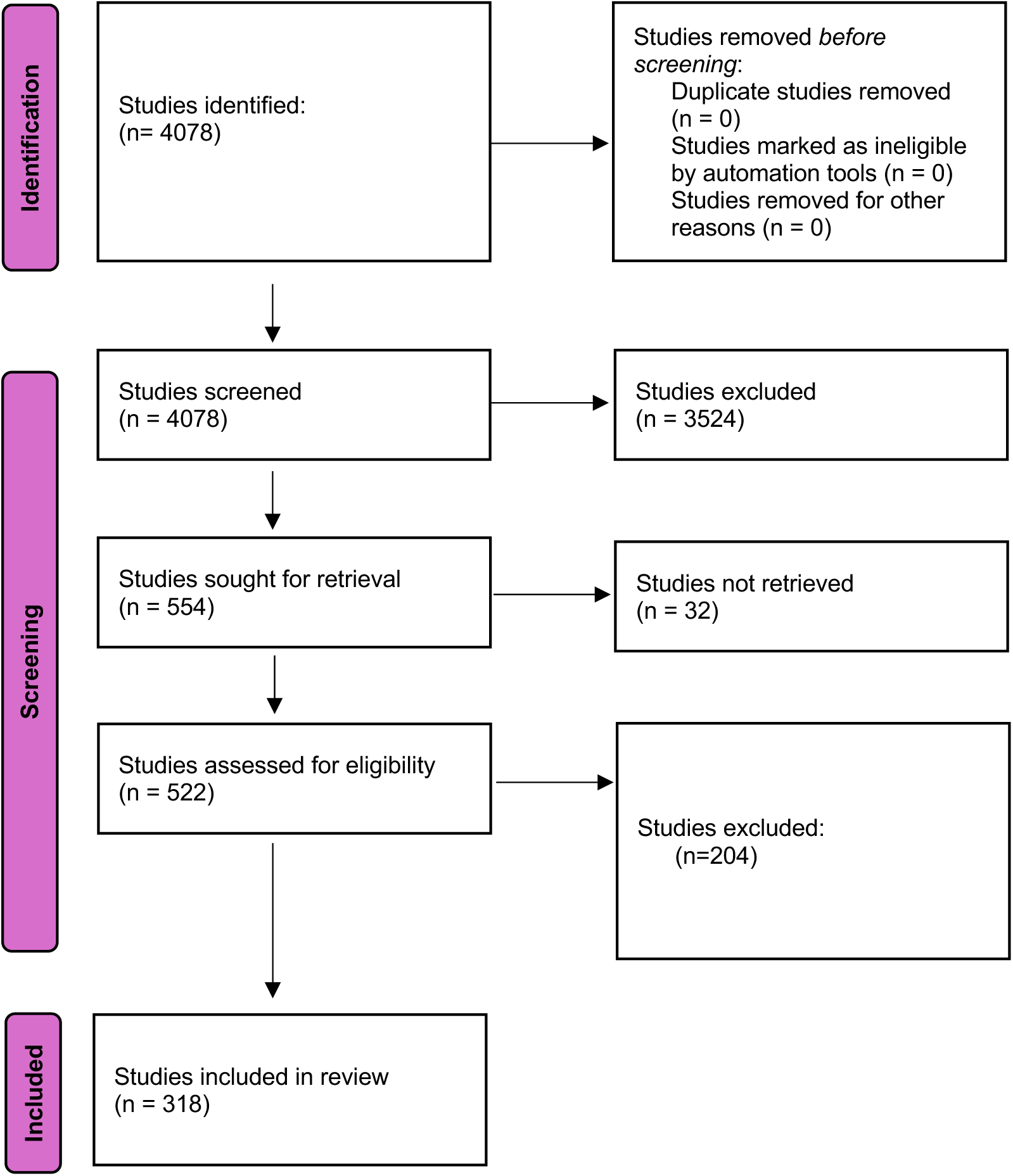
PRISMA flow diagram describing the selection of eligible studies.

### General study characteristics

General study characteristics are presented in Table 1. The most common medical domains that were addressed were oncology (20%), cardiology (16%), and infectious disease (14%). The index test or model studied was a software tool (such as AI or machine learning), in 31% of studies. The median number of study participants was 139 with 25^th^ and 75^th^ percentiles of 68 and 351, respectively, and the number of study participants was below 100 in 32% and above 500 in 17% of the studies.

**Table 1:** General study characteristics for included diagnostic accuracy studies.

| Study characteristic |  | N = 318 |
| --- | --- | --- |
| Medical domain | Oncology | 64 (20%) |
|  | Cardiology | 49 (16%) |
|  | Infectious diseases | 43 (14%) |
|  | Neurology | 41 (13%) |
|  | Pulmonology | 25 (7.9%) |
|  | Other | 94 (30%) |
| Index test was a software tool (e.g., AI) |  | 97 (31%) |
| Number of study participants | median, [P25; P75] | 139 [68; 351] |
|  | <100 | 100 (32%) |
|  | 100-199 | 88 (28%) |
|  | 200-499 | 70 (23%) |
|  | 500+ | 53 (17%) |

### Expert panel characteristics

The number of experts in the panel varied from 1 to 20 (median 3; IQR 2 to 3), with most studies using 3 experts in their panel (46%). In 13% of studies the number of experts in the expert panel was not reported. In over 75% of studies the expert panel consisted of the same experts for all study participants, while in 10% of studies, the study involved a larger pool of experts from which a subset of experts formed each panel. The number of experts involved in a study varied from 1 to 261 (median 3; IQR 2 to 4). Additionally, 12% of studies did not report whether all participants were assessed by the same panel of experts or if panel experts were subsets from a larger group of experts.

The index test or model results were incorporated in the information provided to the expert panel in 18% of the studies. In most cases, these studies were comparative test accuracy studies where the new index test was used as replacement of another test that is used in the current standard of care [11]. For example, one study evaluated whether a new, higher quality CT scan had improved diagnostic performance compared to a currently used CT scan.

In 4% of the studies an expert panel had evaluated the target condition status before the diagnostic accuracy study was conducted, i.e., the data were pre-labeled, e.g., when using a biobank or repository. Thus, most studies set up a new expert panel process to evaluate the target condition status for participants in the study.

At least 8% of the studies did not assess all study participants in the same way, known as differential verification. For example, a study may have used an autopsy to determine target condition presence or absence for patients that have passed away but have used an expert panel to determine target condition status in participants that were alive.

In about 5% of the studies, panel members were asked to indicate how certain they were regarding their judgement on target condition presence. Various methods were used to assess this, including rating on a scale of 1 to 10, describing the level of uncertainty as a percentage, indicating certainty on a diagram, or including multiple categories of judgement (e.g., likely present, likely not present, certainly not present). Some studies asked experts to provide a level of uncertainty to exclude participants that had a high level of uncertainty from further analysis.

**Table 2:**
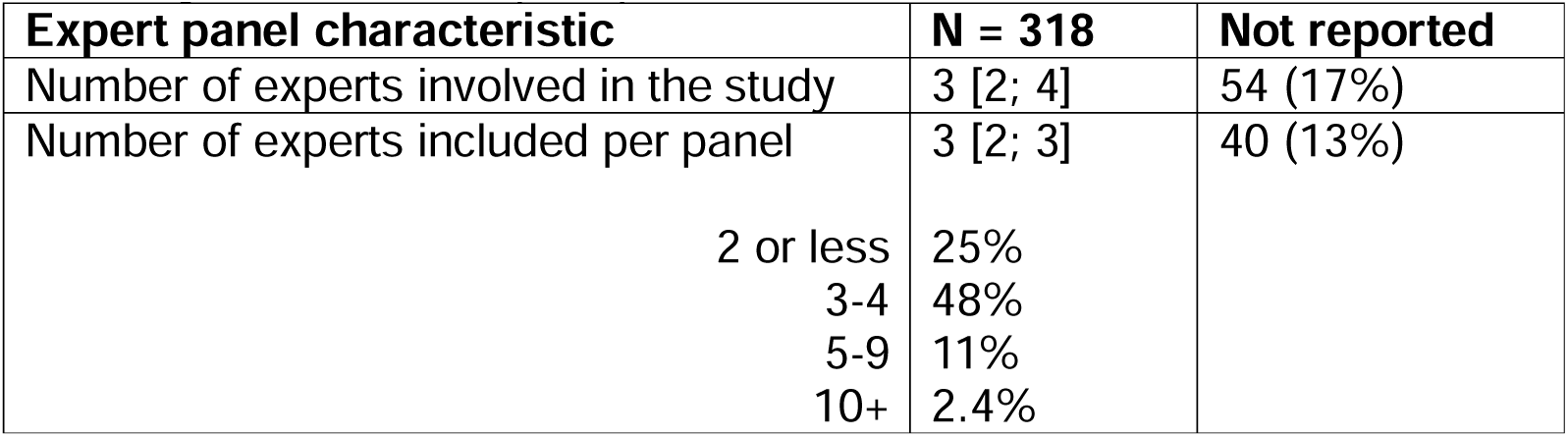
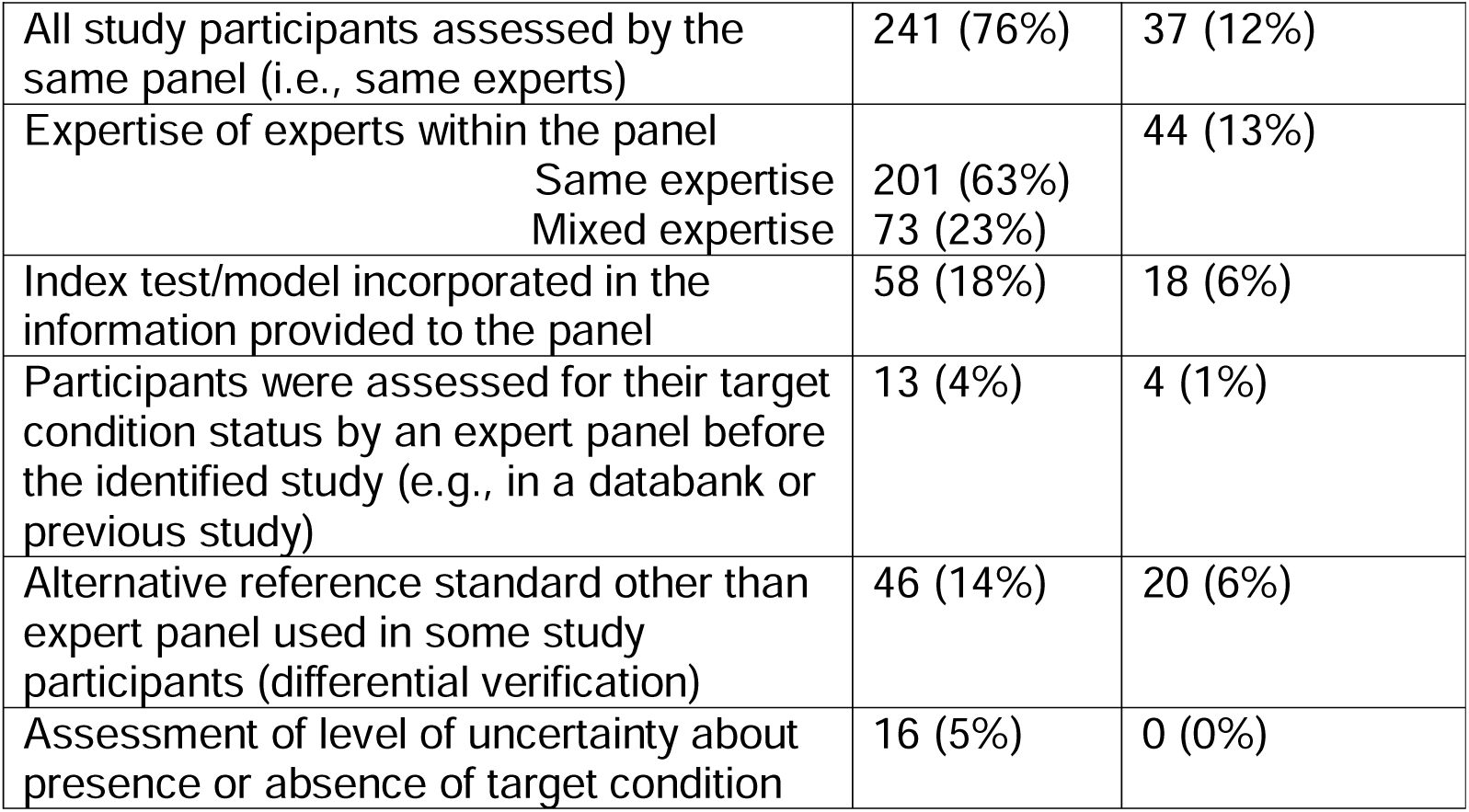
Expert panel characteristics for included studies. Numbers are presented as a total number accompanied by the percentage of the total number of included studies, i.e., n (%), for categorical variables, or median with accompanying interquartile range, i.e., median [p25; p75], for continuous variables.

### Expert panel type

We distinguished four types of expert panels, defined by the method by which they evaluate patient information (step 1), and method by which they decide on the final target condition classification for a given individual (step 2): independent, consensus, staged, and tiebreaker expert panels (see Figure 2). In 20% of included articles, the method by which an expert panel reaches a judgement on target condition status could not be determined.

**Figure 2:**
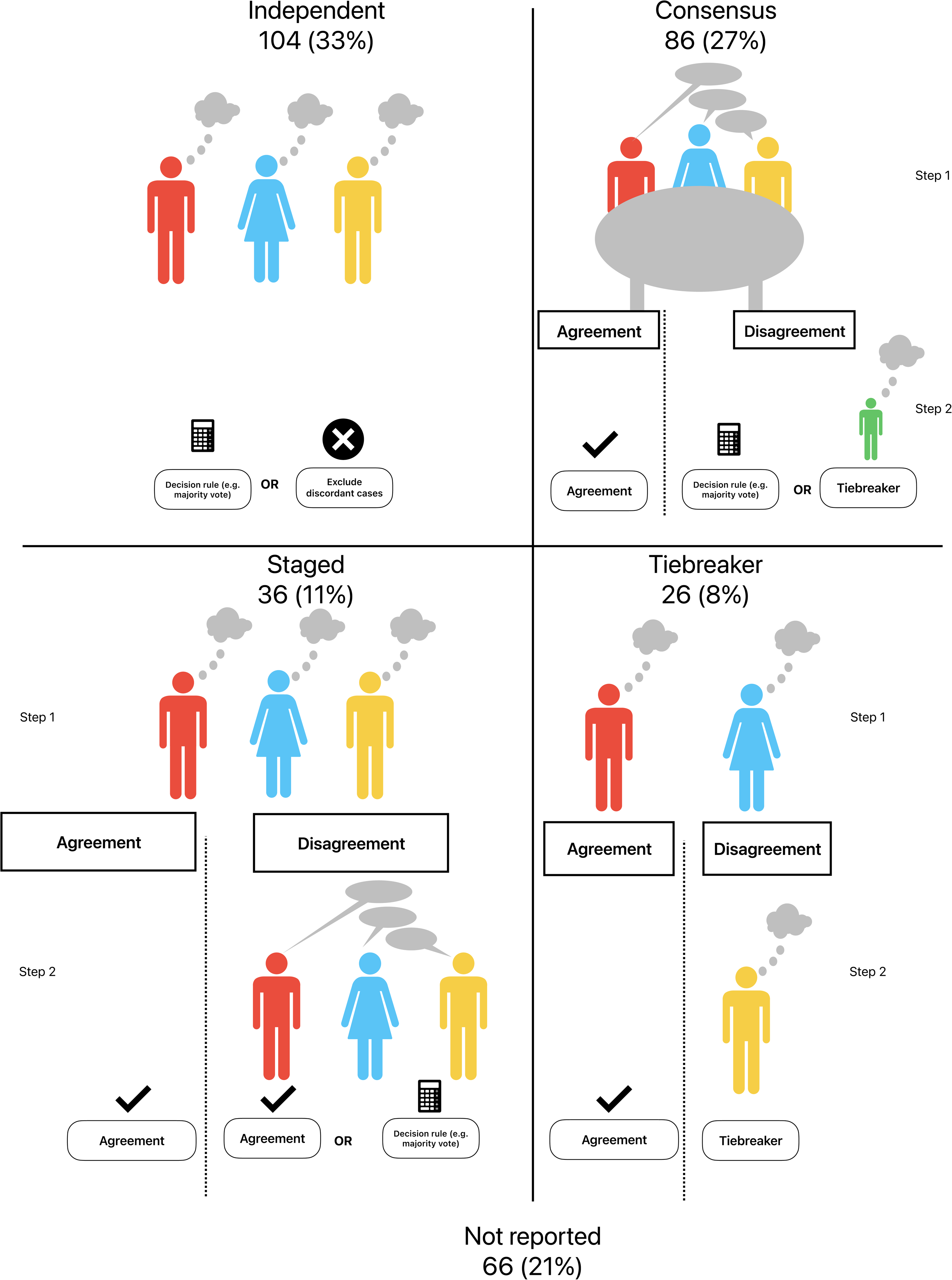
Expert panel types, flow of cases, and decision-making. The number and proportion of studies included in this review using a specific expert panel type are provided.

In the independent expert panel type (n = 104 (33%)), experts are asked to assess patient information independently and provide a decision on target condition status without consulting other experts in the panel. Results were combined using a predefined decision rule (e.g., >50% of experts agree), or by excluding any cases in which experts are not unanimous.

In the consensus expert panel type (n = 86 (27%)), experts discuss each case directly and aim to come to a unanimous agreement on the target condition status for each participant. If there are cases with remaining disagreement, one of the strategies mentioned in the previous section can be used, like a predefined decision rule (majority vote) or calling in a tiebreaker expert.

In the staged expert panel type (n = 36 (11%)), experts are asked to assess patient information independently and provide a decision on target condition status without consulting other experts in the panel. If all experts agree, the target condition status is decided. In case of disagreement, the experts will then jointly discuss the case and attempt to come to a unanimous agreement on the target condition status of each participant.

In the tiebreaker expert panel type (n = 26 (8%)), experts are asked to assess patient information independently and provide a decision on target condition status without consulting other experts in the panel. If the experts agree, the target condition status is defined. If the experts disagree, another expert not involved in the initial assessment is asked to break the tie by providing their assessment, which is then taken as the target condition status.

## Discussion

In this systematic scoping review, we found a large variation in the context, composition, and way that expert panels are used in diagnostic test or model research. Studies used different types of panels and methods for decision-making to provide the final diagnosis for a given participant. Many studies failed to report key characteristics of, and methods used for or by expert panels, complicating replication of their research methodology, as well as assessment of quality, validity or risks of bias.

Besides substantial gaps in reporting quality, several findings stand out. We found that approximately one in five studies incorporate the index test or model into the information provided to experts. The rationale for providing the index test or model results to the expert panel or not was often not provided or discussed. On the one hand, providing the results of the index to the panel could lead to incorporation bias, as the results of the index test under study may become incorrectly used (weighted) in the expert panel judgement, leading to biased accuracy measures [12]. On the other hand, there is the notion that expert panels should receive as much relevant information as possible to allow them to make the most accurate diagnosis. Withholding information on the index test or model results may potentially lead to a less accurate final classification of the target condition status. Which scenario is more likely is difficult to predict, and therefore, in general, a conservative approach is recommended by not providing the result of the test or model under evaluation to the panel. Additional recommendations for the use of expert panels in diagnostic accuracy studies are presented in Box 1.

We also found that some studies excluded participants in whom the expert panel did not agree on the target condition status. We strongly recommend against this, because the accuracy of the index test or model will be overestimated when the hard-to-diagnose cases are excluded.

Collecting information on the remaining uncertainty by the expert panel in each participant was performed in 4% of studies. Measures of remaining uncertainty can provide insight in whether using dichotomous classification is likely to result in biased accuracy estimates and may even be used to more accurately estimate diagnostic test accuracy measures. Methods for this are currently under development but aim to account for the level of uncertainty in the calculation of diagnostic test or model accuracy.

Our results echo those of previous reviews in that there is large variation between studies using expert panels and critical information on expert panel methodology used is often lacking [6]. The independent expert panel type often included only 2 experts and no explanation of the decision-making process in case of disagreement.

Our review has several strengths. It provides an overview of trends in diagnostic test or model studies using expert panels and adds to the literature several aspects of expert panel use that have previously not yet been considered, including incorporation, whether all participants are assessed using the same tests, whether experts were asked to provide their level of uncertainty and distinguishing index tests or models that use software such as AI.

There are also limitations which should be considered. This scoping review which was performed largely systematically is not a comprehensive assessment of expert panels across all literature, but rather is intended to understand trends in studies using expert panels. In contrast to a full systematic review that would search multiple electronic databases, we will not have included every published study on this topic. However, as a systematic scoping review, our study does provide an overview of contemporary use of expert panels in diagnostic test or model accuracy studies and helps to identify directions for further research. Furthermore, this review focused on the use of expert panels in diagnostic test or model studies and as such our results are not necessarily applicable to, for example. prognostic or intervention research or in a clinical setting (i.e., where adjudication panels are used to confirm presence of a clinical outcome). Although certain aspects, e.g., the importance of comprehensive reporting, are also applicable outside a diagnostic accuracy research context.

Several recommendations for future research can be made based on this review. Firstly, there currently is a lack of guidance on the optimal design of expert panel procedures. Research is needed in this regard, thereby considering different criteria, including costs, expert time required to participate in the panel, medical context, general difficulty of the target condition assessment and thus considerations of uncertainty, and performance of the panel as a reference standard. A key issue remains how the performance of an expert panel is influenced by the number of experts in the panel.

Secondly, as big data and artificial intelligence are increasingly used for research, including diagnostic accuracy research, the interplay between these new technologies and expert panels must be assessed in future research.

Finally, we strongly recommend more consistent reporting of the way expert panels are used in diagnostic test or model studies. A detailed description of how the expert panel comes to its final classification is critical information as differences in final classification will directly affect measures of diagnostic accuracy. Complete reporting is therefore required. Our classification can assist researchers in reporting critical information. Studies are difficult to reproduce when information is missing on the index test, the number of experts on the panel and their specialties, the information provided to the panel and whether this includes the index test, or the process by which the expert panel makes their decision. Current reporting guidelines for diagnostic accuracy studies and models, such as STARD and TRIPOD [13, 14], stress the importance of reporting information on the reference standard, but currently do not provide comprehensive guidance for reporting methods specifically used for expert panels, such as decision-making method or choice to incorporate the index test. A specific extension of STARD or TRIPOD will be helpful to authors of diagnostic studies using expert panels. Efforts are already being made to develop a reporting guideline for expert panels [15].

More methodological research and guidance is needed on how to set up an expert panel procedure. The current knowledge is incomplete at best, even for basic components of designing an expert panel such as the impact of the number of experts on the accuracy of the final classification. Another area of interest is collecting and incorporating the remaining uncertainty of experts when estimating measures of accuracy. Also, the link between the expert panel approach and latent class modelling, or using these techniques in conjunction, deserves further attention.

Our review is a clear reminder of the importance of and challenges in obtaining the correct target condition classification in all participants in diagnostic accuracy studies.

### Box 1

Recommendations for expert panels in diagnostic studies

- Consider asking experts to provide a measure of uncertainty and using this measure in your analysis.
- Do not exclude participants if the expert panel disagrees on their target condition status.
- Ensure that the expert panel process is clearly described. Include at least the number of experts in the panel, a description of their expertise, the information made available to the expert panel, a description of the participants evaluated by the expert panel and the decision-making procedure used by the expert panel, including what happens in case of disagreements.
- Follow an appropriate reporting guideline, e.g., STARD or TRIPOD.

## Data Availability

All data and code that support the findings of this study are available at the following URL: https://github.com/BasKellerhuis/Expert-Panel-Reference-Standard-Review.

## Author contributions

BK: Conceptualization, Methodology, Validation, Formal Analysis, Investigation, Data Curation, Writing – Original Draft, Writing – Review and Editing, Visualization, Project Administration. KJ: Conceptualization, Methodology, Supervision, Writing – Review and Editing, Visualization. MK: Validation, Data Curation, Writing – Review and Editing. ES: Supervision, Writing – Review and Editing. LH: Supervision, Writing – Review and Editing. KM: Supervision, Writing – Review and Editing. JR: Conceptualization, Methodology, Supervision, Writing – Review and Editing.

## Acknowledgements

We would like to thank Rene Spijker (Cochrane Netherlands) for his expertise as an information specialist and help with the systematic search.

## Appendix A

Search conducted on the 6^th^ of October 2022 in PubMed for records published between the 1^st^ of June 2012 and the 1^st^ of October 2022 using the following search string:

**(diagnosis OR diagnostic*) AND ((panel AND expert*) OR (consensus AND (panel OR opinion OR expert*))) AND (“diagnostic accuracy” OR sensitivity OR specificity OR AUC OR “c-statistic” OR “predictive value” OR PPV OR NPV OR ROC) NOT ((“systematic review”[Publication Type]) OR (“review”[Publication Type]))**

## Appendix B

| *Category* | *Explanation* | *Expected result* |
| --- | --- | --- |
| Target condition | The target condition(s) or health status that is being diagnosed in the study | Name of target condition |
| Number of participants | Number of evaluations made by expert panel | Number |
| Expert panel type | Independent (each expert makes a classification), Tiebreaker (e.g. two experts make a classification, if they don't agree a third expert breaks the tie), consensus (classification made by discussion or consensus), staged approach (individual with tiebreaker by consensus) | One of: **independent, tiebreaker, consensus, staged** |
| Number of experts in study | Total number of experts that have made at least one decision/estimate/classification | Number |
| Number of experts per panel | Number of experts involved in each decision per participant | Number |
| Specialties | Specialties experts belong to (e.g. 2 radiologists, 1 cardiologist) NB for each panel, not the total number of experts | List of specialties |
| Experts are the same in all decisions | Experts within the expert panel are the same for each patient (e.g. 3 experts in total and all 3 evaluate each participant would be a Yes) | Yes/no |
| Differential verification | Reference standard is different for different participants (e.g. some patients receive an expert panel and others a biopsy OR some patients receive a decision by a different number of experts or different specialties than others) | Yes/no |
| Component tests | Description of the information a panel has access to when making a decision (e.g. patient history and characteristics, biomarkers, tests) | List of tests |
| Incorporation | Index test (the test under review) is one of the component tests | Yes/no |
| Machine learning / AI | Study is about machine learning/AI or about neural networks/deep learning (if authors do not use ML/AI/NN/DL/Automated/algorithm/software to describe their work then 'no') | Yes/no |
| Pre-labeled data | Expert panel was originally part of another study or initiative | Yes/no |
| Outcome | How was the expert panel asked to record their judgement? In particular, were they asked to provide a measure of uncertainty? | Dichotomous, categorical, other (e.g., confidence, percentages)<br>[multiple different diagnoses was not |
|  |  | considered<br>categorical,<br>categorical here<br>means levels of<br>confidence] |

